# Repurposing of drugs for Covid-19: a systematic review and meta-analysis

**DOI:** 10.1101/2020.06.07.20124677

**Authors:** Pinky Kotecha, Alexander Light, Enrico Checcucci, Daniele Amparore, Cristian Fiori, Francesco Porpiglia, Prokar Dasgupta, Oussama Elhage

## Abstract

**Objective:** The aim of this systematic review is to evaluate the data currently available regarding the repurposing of different drugs for Covid-19 treatment. Participants with suspected or diagnosed Covid-19 will be included. The interventions being considered are drugs being repurposed, and comparators will include standard of care treatment or placebo.

**Methods:** We searched Ovid-MEDLINE, EMBASE, Cochrane library, clinical trial registration site in the UK(NIHR), Europe (clinicaltrialsregister.eu), US (ClinicalTrials.gov) and internationally (isrctn.com), and reviewed the reference lists of articles for eligible articles published up to April 22, 2020. All studies in English that evaluated the efficacy of the listed drugs were included. Cochrane RoB 2.0 and ROBINS-I tool were used to assess study quality. This systematic review adheres to the PRISMA guidelines. The protocol is available at PROSPERO (CRD42020180915).

**Results:** From 708 identified studies or clinical trials, 16 studies and 16 case reports met our eligibility criteria. Of these, 6 were randomized controlled trials (763 patients), 7 cohort studies (321 patients) and 3 case series (191 patients). Chloroquine (CQ) had a 100% discharge rate compared to 50% with lopinavir-ritonavir at day 14, however a trial has recommended against a high dosage due to cardiotoxic events. Hydroxychloroquine (HCQ) has shown no significant improvement in negative seroconversion rate which is also seen in our meta-analysis (p=0.68). Adverse events with HCQ have a significant difference compared to the control group (p=0.001). Lopinavir-ritonavir has shown no improvement in time to clinical improvement which is seen in our meta-analyses (p=0.1). Remdesivir has shown no significant improvement in time to clinical improvement but this trial had insufficient power.

**Discussion:** Due to the paucity in evidence, it is difficult to establish the efficacy of these drugs in the treatment of Covid-19 as currently there is no significant clinical effectiveness of the repurposed drugs. Further large clinical trials are required to achieve more reliable findings. A risk-benefit analysis is required on an individual basis to weigh out the potential improvement in clinical outcome and viral load reduction compared to the risks of the adverse events. (1-16)

## Introduction

Starting in December 2019, there was a pneumonia outbreak of unknown cause in Wuhan, Hubei province of China (17). The origin of the virus is unknown but there is an epidemiological link the Huanan Seafood Wholesale Market where there was a sale of wild animals, such as bats (17). After notification of the World Health Organisation (WHO) on 31 December 2019, scientists were able to isolate a 2019-nCoV from a patient and subsequently perform genome sequencing by the 7^th^ of January. Since then many cases have emerged internationally leading to the WHO declaring the novel 2019-nCoV (Covid-19) outbreak a global pandemic (18).

Similar clinical features to previous betacoronavirus infections have been noted, including presentations with fever, dry cough and dyspnoea but very few presentations with prominent upper respiratory tract signs and symptoms such as rhinorrhoea, sneezing or sore throat (19). On imaging, bilateral ground-glass opacities on chest computed tomography (CT) scans have been noted. The patients with severe illness developed Acute Respiratory Distress Syndrome (ARDS) and required Intensive Care Unit (ICU) admission and oxygen therapy. These features bear resemblances with the severe acute respiratory distress syndrome (SARS-CoV) and Middle East respiratory syndrome coronavirus (MERS-CoV) infections. However, Covid-19 patients rarely develop intestinal symptoms such as diarrhoea which was present in about 20-25% of those with SARS-CoV and MERS-CoV. The mortality rate has been similarly described by cohorts (19-22) as 4-15%.

With many describing the memories of the novel coronavirus outbreak in China, SARS-CoV in 2003 (17), drugs used during SARS-CoV and Middle East respiratory syndrome coronavirus (MERS-CoV) are being considered (19).

With no current universally agreed treatment for Covid-19, the current care advised is for supportive management depending on patient’s needs, including antipyretics for fever and oxygen therapy.

The repurposing of drugs can provide an avenue to find treatment options for Covid-19 which has currently infected over 3.2 million people as of 2 May 2020 reported by the WHO (23).

Hydroxychloroquine (HCQ) and chloroquine (CQ), both anti-malarial drugs, have been authorised by both US and French authorities as there is no adequate approved and available alternative to treat Covid-19 (24). These drugs have been shown to have potent anti-viral activity against Covid-19 in in-vitro studies (25, 26). Both can have adverse effects with HCQ, a derivative of CQ, being less toxic when used long term. Therefore, recently, high-dosage of CQ (12g) with either azithromycin and oseltamir has not been recommended in patients with severe Covid-19 due to safety issues (27).

The use of anti-virals has also been trialled as previously, screening of approved drugs identified anti-virals to have an inhibitory activity on SARS-CoV. Lopinavir, an anti-viral, also has activity against MERS-CoV both in vitro and in animal models ^14^. Therefore, due to the homogeneity of SARS-CoV-2 compared to the mentioned zoonotic viruses, anti-virals which have previously been used are being repurposed. Remdesivir has also demonstrated effective control of Covid-19 in-vitro (26) and has since been authorised for emergency use by the Food and Drug Administration (FDA) (28).

The aim of this systematic review is to evaluate the data currently available regarding the repurposing of different drugs for Covid-19 treatment.

## Methods

### Search strategy and selection criteria

This systematic review adheres to the Preferred Reporting Items for Systematic Reviews and Meta-analyses (PRISMA) guidelines (29). We searched Ovid-MEDLINE, EMBASE, Cochrane library for articles published any time up to April 22, 2020. We also searched clinical trial registration sites in the UK (NIHR), Europe (clinicaltrialsregister.eu), US (ClinicalTrials.gov) and internationally (isrctn.com). We examined the reference lists of articles to identify additional studies.

The following search term was used: (favipiravir or remdesivir or galidesivir or ivermectin or oseltamivir or ganciclovir or lopinavir or ritonavir or darunavir or CQ or HCQ or arbidol or azithromycin or amoxicillin or moxifloxacin or ceftriaxone or antifungals or androgen receptor blockers or tea or traditional Chinese medicine) and (“2019 nCoV” or 2019nCoV or “2019 novel coronavirus” or “COVID 19” or COVID19 or “new coronavirus” or “novel coronavirus” or “SARS CoV-2” or (Wuhan coronavirus) or “COVID 19” or “SARS-CoV” or “2019-nCoV” or “SARS-CoV-2”)

Only studies in English that evaluated the efficacy of the listed drugs were included. This included randomized controlled trials (RCTs) as well as observational studies (including cohort and control studies). Case reports were also collated.

Patients with suspected and consequently diagnosed with Novel coronavirus (Covid-19) will be included, with the following interventions: favipiravir, remdesivir, galidesivir, ivermectin, oseltamivir, ganciclovir, lopinavir/ ritonavir, darunavir, chloroquine, hydroxychloroquine, arbidol, azithromycin, amoxicillin, moxifloxacin, ceftriaxone, antifungals, androgen receptor blockers, tea and traditional Chinese medicine will be considered. Comparators will include standard of care treatment or placebo. The main outcomes will be time to clinical recovery, benefits in reducing mortality and reduction in viral load.

We excluded studies in other languages when no translation was available, review articles, commentaries and letters to editors.

One reviewer (PK) extracted data using a spreadsheet and a second (AL) validated data extraction. Descriptive and quantitative data were entered into a spreadsheet.

The protocol is available at PROSPERO (CRD42020180915).

### Data analysis

One author (PK) extracted the data which was confirmed by another author (AL). Duplicate studies and clinical trials were removed as shown in PRISMA diagram (Figure 1).

**Figure 1:**
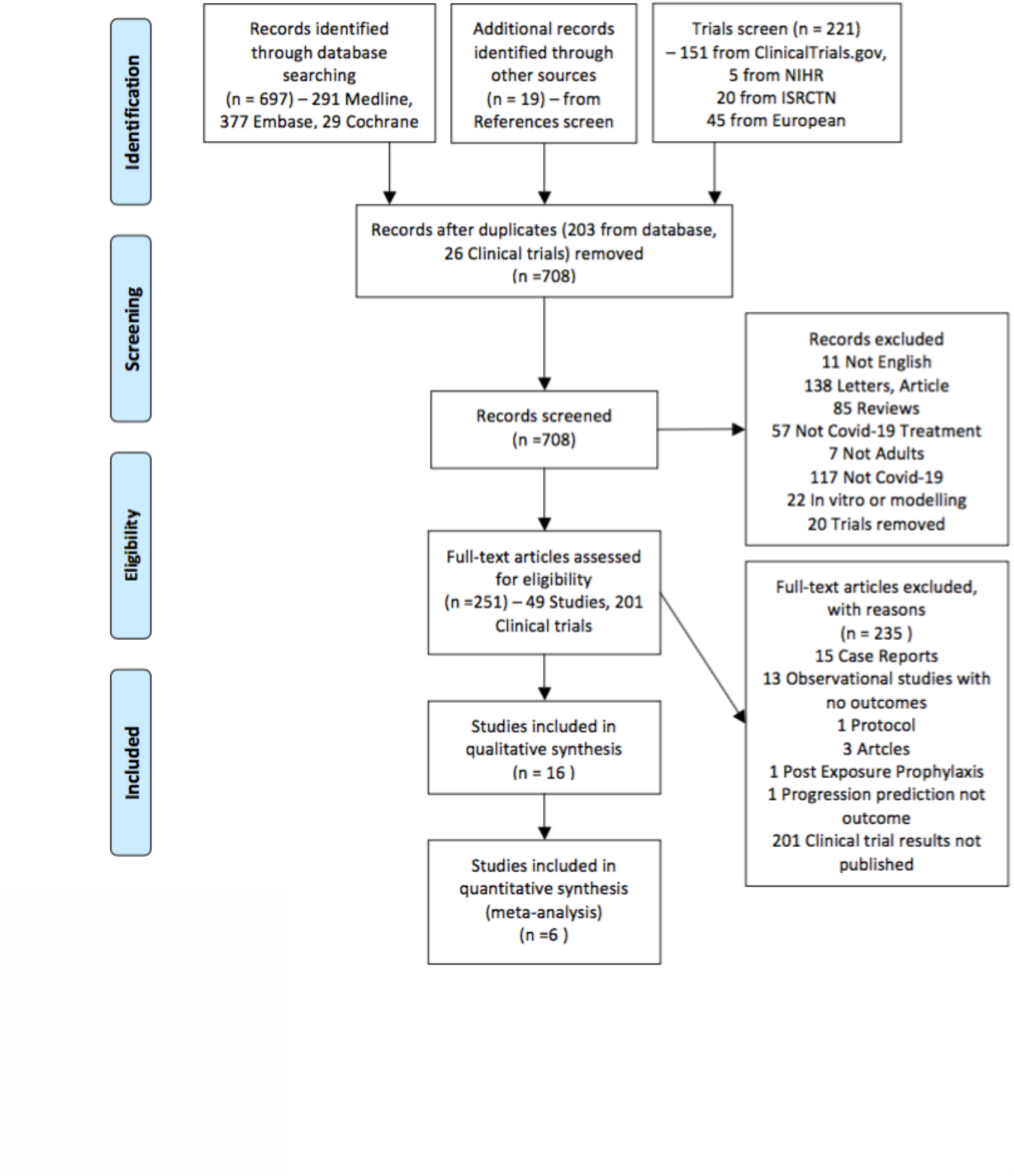
PRISMA diagram – study selection (29)

Main summary measures

- Time to clinical recovery
- Benefits in reducing mortality
- Reduction in viral load
- Measures of effect as appropriate for the studies in question – hazards ratios, odds ratios and mean difference

Preplanned secondary outcomes included: Negative Seroconversion, time to a negative Covid-19 seroconversion, time to discharge, symptom alleviation, changes in blood tests, lung function, rate of respiratory failure, oxygen therapy requirement, non invasive ventilation requirement, radiological results, all cause mortality, rate of patients needing intensive care, Length of hospital stay, overall survival and adverse events.

Quality of studies (risk of bias) was assessed using the Cochrane RoB 2.0 for RCTs (Figure 2) and ROBINS-I (30) for non-RCTs (Figure 3).

**Figure 2:**
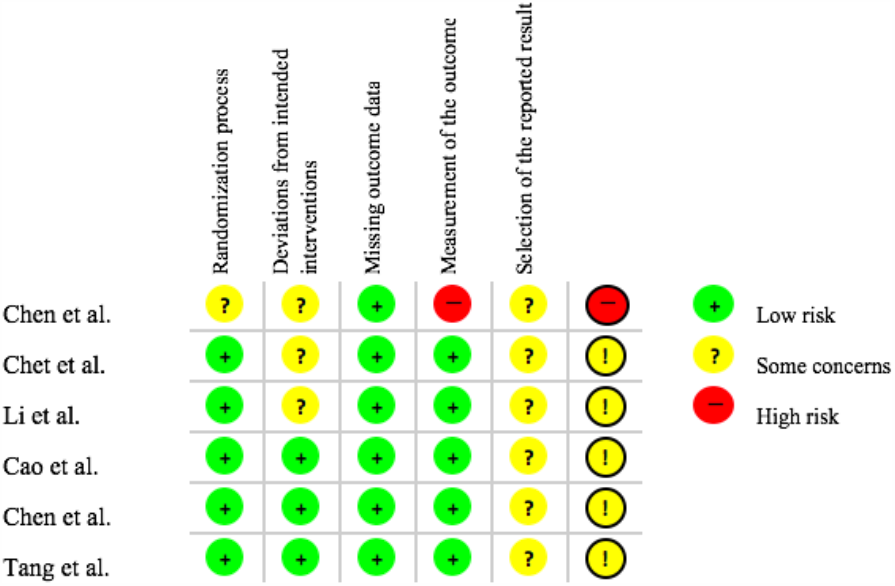
Risk of Bias for Randomised Controlled Trials using RoB 2.0 tool.

**Figure 3:**
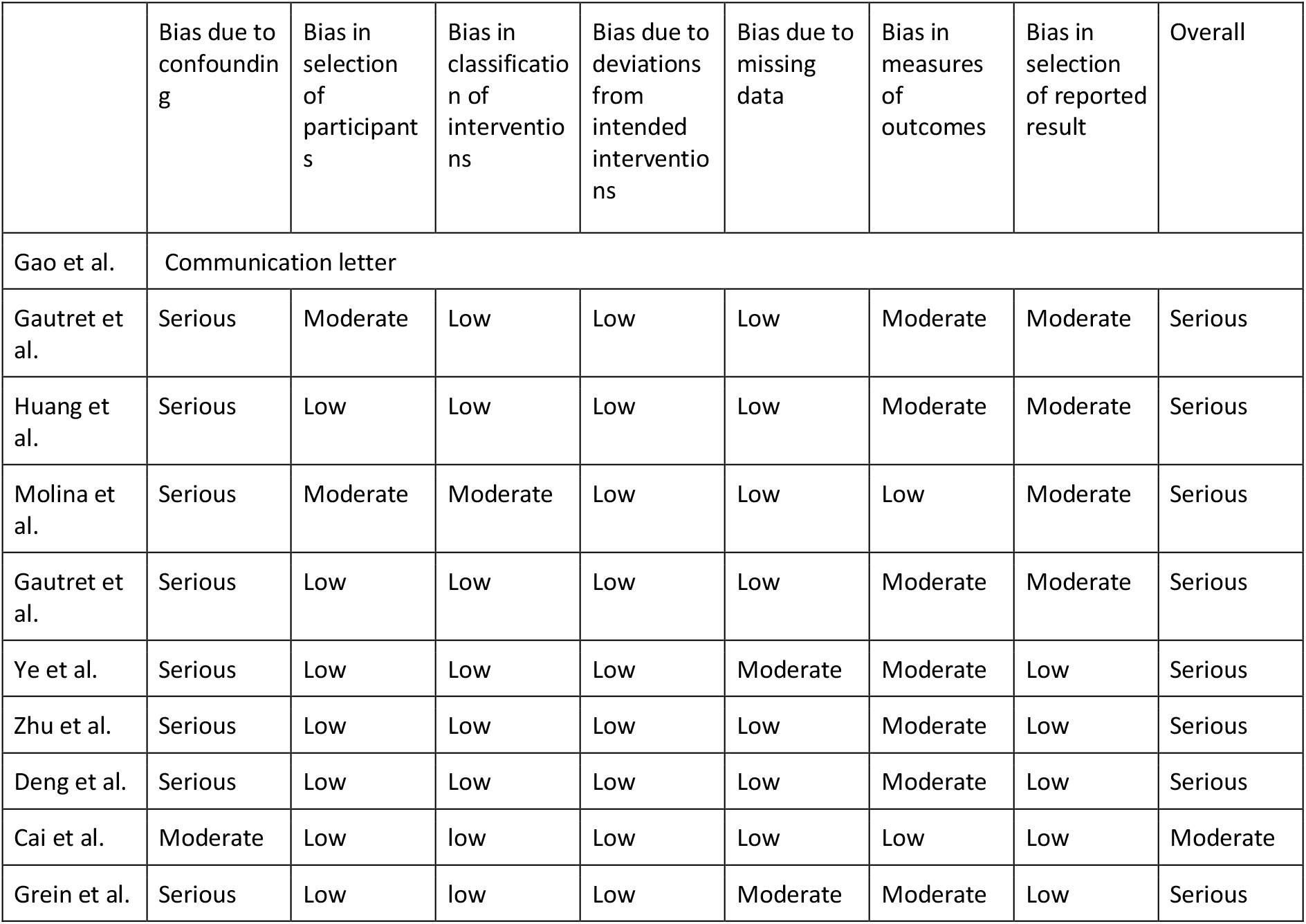
Risk of Bias Assessment for Non-randomised Controlled trials using ROBINS-I tool (30)

Where trials examined similar outcomes and were considered suitably homogenous, meta-analysis was performed. For adverse events and rates of negative seroconversion, odds ratios with 95% confidence intervals were calculated. The chi squared test was used to assess for heterogeneity. If p<0.10 for heterogeneity, then a random effects model was used, otherwise a fixed effects model was used. All meta-analyses were performed using the Cochrane Review Manager (v.5.3, 2014; Cochrane Initiative).

## Results

## Overview

Our literature search identified 937 studies or clinical trials, with 708 included after removal of duplicates. After abstract screening, 251 were included for full-text screening and of these 16 were included in qualitative synthesis (Figure 1), with 6 of these being randomized controlled trials.

Characteristics of studies (1-16) included are summarized in Table 1. 12 of the studies were carried out in China. The eligibility criteria varied, with some studies including any adult with Covid-19 while others restricted to mild, moderate or severe illness, defined differently by studies (summarised in Table 1). In total, from 16 studies, a total of 1275 patients were included in this systematic review. Of these, 141 received HCQ, 110 CQ, 91 HCQ + azithromycin, 283 Lopinavir-ritonavir (LPV-r), 171 arbidol, 16 arbidol and LPV/e, 151 favipavir, 53 remdesivir and 259 standard care. Standard care included, as necessary, supplemental oxygen, noninvasive and invasive ventilation, antibiotic agents, vasopressor support, renal replacement therapy, and extracorporeal membrane oxygenation (ECMO).

**Table 1:** Characteristics of study (* - no dose reported, HCQ – hydroxychloroquine, LPV-r – lopinavir-ritonavir)

| Author | Study Type | Country | Dates of study | Inclusion Criteria | Follow up period | Total number of patients | Number of patients in each arm | Age Median (IQR) | Male | Intervention | Outcomes |
| --- | --- | --- | --- | --- | --- | --- | --- | --- | --- | --- | --- |
| Hydroxychloroquine and chloroquine |  |  |  |  |  |  |  |  |  |  |  |
| Chen et al. (1) | RCT | China | NR | NR | NR | 30 | 15 | NR | NR | HCQ 400mg OD for 5 days | Negative conversion rate of Covid-19 nucleic acid in respiratory pharyngeal swab on days 7 |
|  |  |  |  |  |  |  | 15 | NR | NR | Conventional treatment |  |
| Chen et al. (2) | RCT | China | 4 - 28 February 2020 | Age ≥18 years with RT-PCR-confirmed Covid-19, CT-proven pneumonia, and mild illness (SaO2 >93%) | 5 days | 62 | 31 | 44.1 ± 16.1 | 14 (45.2%) | 5 day HCQ 400mg/day | Time to clinical recovery (TTCR) - return of body temperature and cough relief, maintained for >72h- Body temperature <36.5°C on surface, <37.2°C under armpit and mouth or <37.8°C in rectum and tympanic membrane. Clinical characteristics. Radiological changes |
|  |  |  |  |  |  |  | 31 | 45.2 ± 15.3 | 15 (48.3%) | Control |  |
| Gao et al. (3) | Case Series | China | NR | NR | NR | >100 | N/A | NR | NR | Chloroquine Phosphate * | NR |
| Gautret et al. (4) | Case Series | France | 3 to 21 March 2020 | RT-PCR-confirmed Covid-19 | At least 6 days | 80 | N/A | 52 (18-88) | 43 (53.8%) | HCQ and azithromycin * | Clinical outcome, contagiousness (PCT + culture), length of stay |
| Huang et al. (5) | Cohort | China | 27 January to 15 | Age ≥18 years with RT-PCR-confirmed Covid-19 | 14 days | 22 | 10 | 41.5 (33.8–50.0) | 7 (30%) | Chloroquine * | Real Time PCR for viral RNA, lung CT for improvement, length of stay (discharge |
|  |  |  | February 2020 |  |  |  | 12 | 53.0 (41.8–63.5) | 6 (50%) | LPV-r | criteria: temperature returned to <36.6°C in axilla for >3days, respiratory symptoms improved, pulmonary imaging showing absorption, detection of respiratory pathogenic nucleic acid negative twice in a row) |
| Molina et al. (6) | Case Series | France | NR | Hospitalised | 5-6 days | 11 | N/A | 58.7 (range 20-77) | 7 (63.6%) | HCQ 600mg/day for 10 days and Azithromycin (500mg day1 and 250mg days2-5) | Viral clearance, clinical outcome |
| Gautret et al. (7) | Cohort | France | “Early” March to 16 March 2020 | Age >12 years with RT-PCR-confirmed Covid-19 | 14 days | 36 | 20 | 51.2 ± 18.7 | 9 (45%) | HCQ 200mg TDS 10 days | Viral clearance , clinical outcome |
|  |  |  |  |  |  |  | 16 | 37.3 ± 24.0 | 6 (37.5%) | Control |  |
| Tang et al. (8) | RCT | China | 11 - 29 February 2020 | Age ≥18 years with RT-PCR-confirmed Covid-19 | 28 days | 150 | 75 | 46 | 55% | HCQ 1200mg for 3 days. 800mg daily thereafter. Total duration - 2 or 3 weeks for mild/moderate or severe patients respectively | Negative seroconversion within 28-day, clinical outcome |
|  |  |  |  |  |  |  | 75 |  |  | Control |  |
| Anti-virals |  |  |  |  |  |  |  |  |  |  |  |
| Ye et al. (9) | Cohort | China | 22 - 29 January | RT-PCR-confirmed Covid-19 | 10 days | 47 | 42 |  | 21 (50%) | LVP 400mg- r 100mg BD | Body temperature, blood results |
|  |  |  |  |  |  |  | 5 |  | 1 (20%) | Control |  |
| Zhu et al. (10) | Cohort | China | 23 January - 29 February 2020 | Covid-19 as diagnosed by the Chinese guideline | 14 days | 50 | 34 | 40.5 (34.8 – 52.3) | 20 (58.8%) | LPV *- r* | RT-PCR, fever duration |
|  |  |  |  |  |  |  | 16 | 26.5 (23.3 – 52.5) | 6 (37.5%) | Arbidol * |  |
| Li et al. (11) | RCT | China | NR | Age 18-80 years with RT-PCR-confirmed Covid-19, which was mild (mild symptoms but no signs of pneumonia on imaging) or moderate (fever, respiratory symptoms and pneumonia on imaging) | 14 days | 86 | 34 |  |  | LPV 200mg - r 50mg BD (500mg) | Average time of positive to negative conversion. Conversion rates at day 7 and 14, fever resolution, radiological changes, clinical outcome |
|  |  |  |  |  |  |  | 35 |  |  | Arbidol 200mg TDS |  |
|  |  |  |  |  |  |  | 17 |  |  | No antiviral |  |
| Deng et al. (12) | Cohort | China | 17 January - 13 February 2020 | Age ≥18 years with RT-PCR-confirmed Covid-19 pneumonia without invasive or non-invasive ventilation | 14 days | 33 | 16 | 41.8(14.08) | 7 (43.8%) | Arbidol and LPV-r | Negative conversion rate, radiological changes |
|  |  |  |  |  |  |  | 17 | 47.25(17.25) | 10 (58.8%) | LPV-r |  |
| Cai et al. (13) | Cohort | China | 30 January - 14 February 202 | Age 16-75 years, RT-PCR-confirmed Covid-197 day duration from disease onset, not severe clinical condition (RR>30, | 14 days | 80 | 35 | 43 (35.5-59) | 14 (40%) | Favipiravir day 1 1600mg BD, day 2-14 600mg BD | Time of viral clearance, radiological changes by Day 14 |
|  |  |  |  | SaO2<93%, respiratory failure, shock, multi-organ failure requiring ICU) |  |  | 45 | 49 (36–61) | 21 (46.7%) | Control, LPV- r 400mg/100mg BD |  |
| Cao et al. (14) | RCT | China | 18 January - 3 February 2020 | Age ≥18 years with RT-PCR and radiologically-confirmed Covid-19 that was severe (SaO2<94% on room air or PaO2:FiO2 <300mgHg) | 14 days | 199 | 99 (94 received) | 58 (49-68) | 60.3% | LPV-e 400mg and 100mg PO BD | Time to clinical improvement: time from randomisation to an improvement of 2 points on a seven-category ordinal scale or live discharge from hospital |
|  |  |  |  |  |  |  | 100 |  |  | Standard Care |  |
| Chen et al. (15) | RCT | China | 20 February - 1 March 2020 | Age ≥18 years with evidence of Covid-19 clinically, radiologically, or lymphopenia | 7 days | 236 | 116 | 29 (25.00%) were ≥65 years | 59 (50.86%) | Favipavir 1600mg BD/ first day, 600mg BD for 10 days | Clinical recovery rate at 7 days from beginning of treatment - >72h recovery of body (axillary) temperature <36.6°C, respiratory rate, SaO2 >98% and cough relief (mild or none) |
|  |  |  |  |  |  |  | 120 | 41 (34.17%) | 51 (42.5%) | Arbidol 200mg TDS |  |
|  |  |  |  |  |  |  |  | were ≥65 years |  |  |  |
| Grein et al. (16) | Cohort | US, Europe, Canada, Japan | 25 January - 30 March 2020 | RT-PCR-confirmed Covid-19 with SaO2<94% on air or a need for oxygen support | 28 days | 53 |  | 64 (48-71) | 40 (75%) | Remdesivir | Clinical improvement: live discharge from hospital or decrease of at least 2 points from baseline on modified ordinal six point scale |

Primary outcome of the studies included are also variables including time to clinical recovery and viral clearance. Clinical improvement data for individual studies is summarized in Table 2. Data on Virology and Radiology is shown in Table 3.

**Table 2:** Clinical Improvement (HCQ: hydroxychloroquine, LPV-r: Lopinavir-ritonavir)

| Author | Intervention | Clinical Recovery | Median days for body temperature normalisation | O2 or NIV therapy | Duration of Invasive mechanical ventilation | ICU - rate of patients, length of stay | Length of stay in hospital | Time to discharge | Mortality |
| --- | --- | --- | --- | --- | --- | --- | --- | --- | --- |
| **Hydroxychloroquine and chloroquine** |  |  |  |  |  |  |  |  |  |
| Chen et al. (1) | HCQ |  | 1 (0-2) |  |  |  |  |  |  |
|  | Conventional treatment |  | 1 (0-3) |  |  |  |  |  |  |
| Chen et al. (2) | HCQ |  | 3.2 (1.3) |  |  |  |  |  |  |
|  | Control |  | 2.2 (0.4) |  |  |  |  |  |  |
| Gautret et al. (4) | HCQ and azithromycin |  |  | O2: 12 (15%) |  | 3 (3.8%) | 4.6 ± 2.1 (1-11) | 4.1 ± 4.2 (1-10) | 1 (1.2%) |
| Huang et al. (5) | Chloroquine |  |  |  |  |  |  | By day 14: 100% discharged |  |
|  | LPV-r |  |  |  |  |  |  | By day 14: 50% discharged |  |
| Molina et al. (6) | HCQ and azithromycin |  |  |  |  | 2 (18.2%) |  |  | 1 (9.1%) |
| Tang et al. (8) | HCQ | Median time to symptom alleviation: 19 days |  |  |  |  |  |  |  |
|  | Control | Median time to symptom alleviation: 21 days |  |  |  |  |  |  |  |
| **Anti-virals** |  |  |  |  |  |  |  |  |  |
| Ye et al. (9) | LPV-r |  | 4.8 ± 1.94 days (p=0.0364) |  |  |  |  |  |  |
|  | Control |  | 7.3 ± 1.53 days |  |  |  |  |  |  |
| Cao et al. (14) | LPV-r | 16 |  | O2 requirement: 12 (9-16) | 4 (3-7) | Average stay in ICU - 6 | 14 (12-17) | 12 | 28 day mortality: 19 (19.2%).<br>9 days to death (6-13) |
|  |  | Day 7: 6 (6.1%)<br>Day 14: 45 (45.4%)<br>Day 28: 78 (78.8%) |  |  |  |  |  |  |  |
|  | Standard Care | 16 |  | O2 requirement - 13 (6-16) days | 5 (3-9) | Average stay in ICU: 11 | 16 (13-18) | 14 | 28 day mortality: 25 (25.0%).<br>12 days to death (6-15) |
|  |  | Day 7: 2 (2%)<br>Day 14: 30 (30%)<br>Day 28: 70 (70%) |  |  |  |  |  |  |  |
| Chen et al. (15) | Favipavir | Day 7: 71 (61.2%) |  | O2 + NIV – 21 patients (18.1%) |  |  |  |  | 0 |
|  | Arbidol | Day 7: 62 (51.7%) |  | O2 + NIV – 27 patients (22.5%) |  |  |  |  | 0 |
| Grein et al. (16) | Remdesivir | Day 28: 84% improvement |  | 36 (68%) improvement in category of O2 support<br>8 (15%) worsening |  |  |  |  | 0.56 deaths per 100 hospitalisation days. 7 (13%) deaths<br>15 days to death |

**Table 3:**
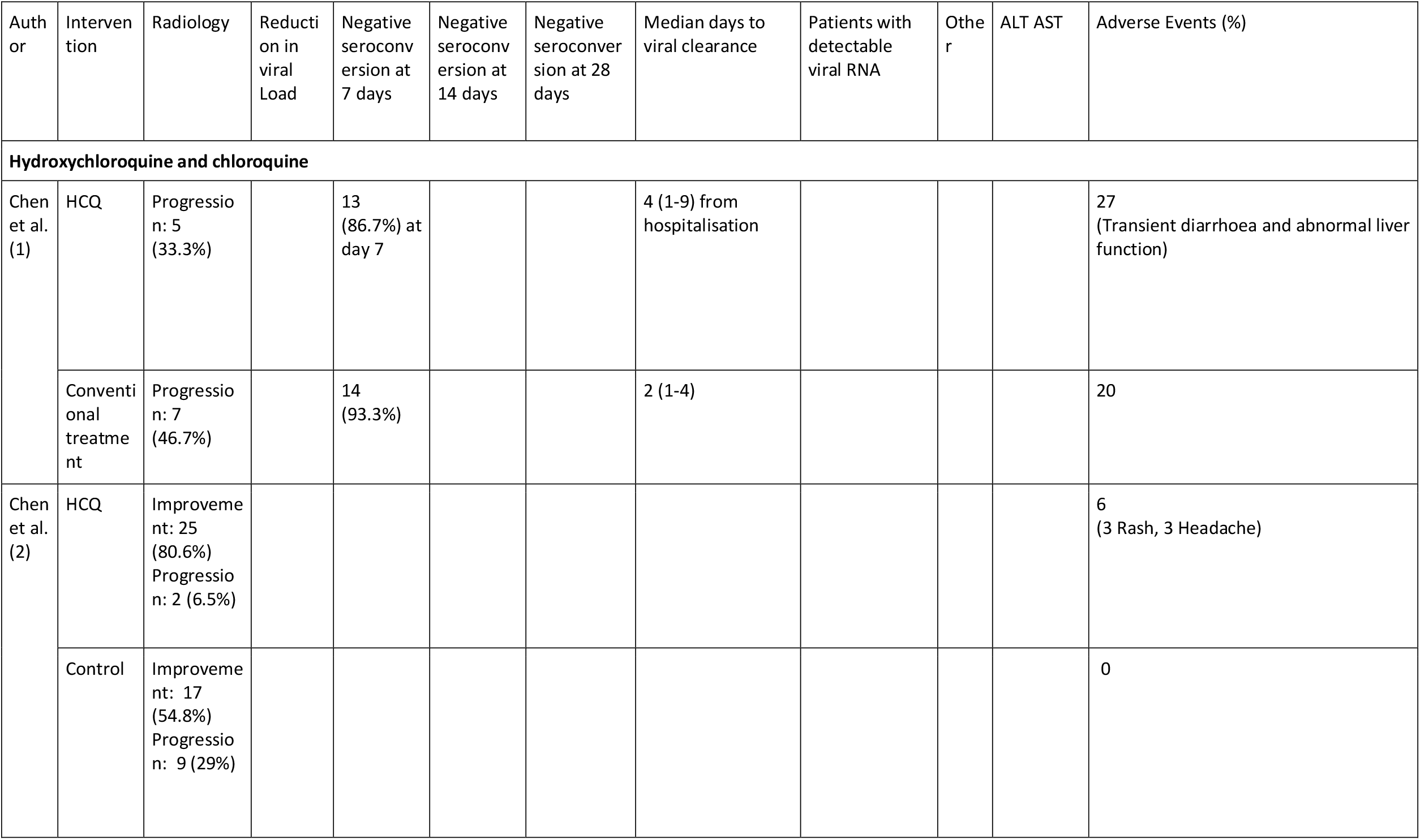
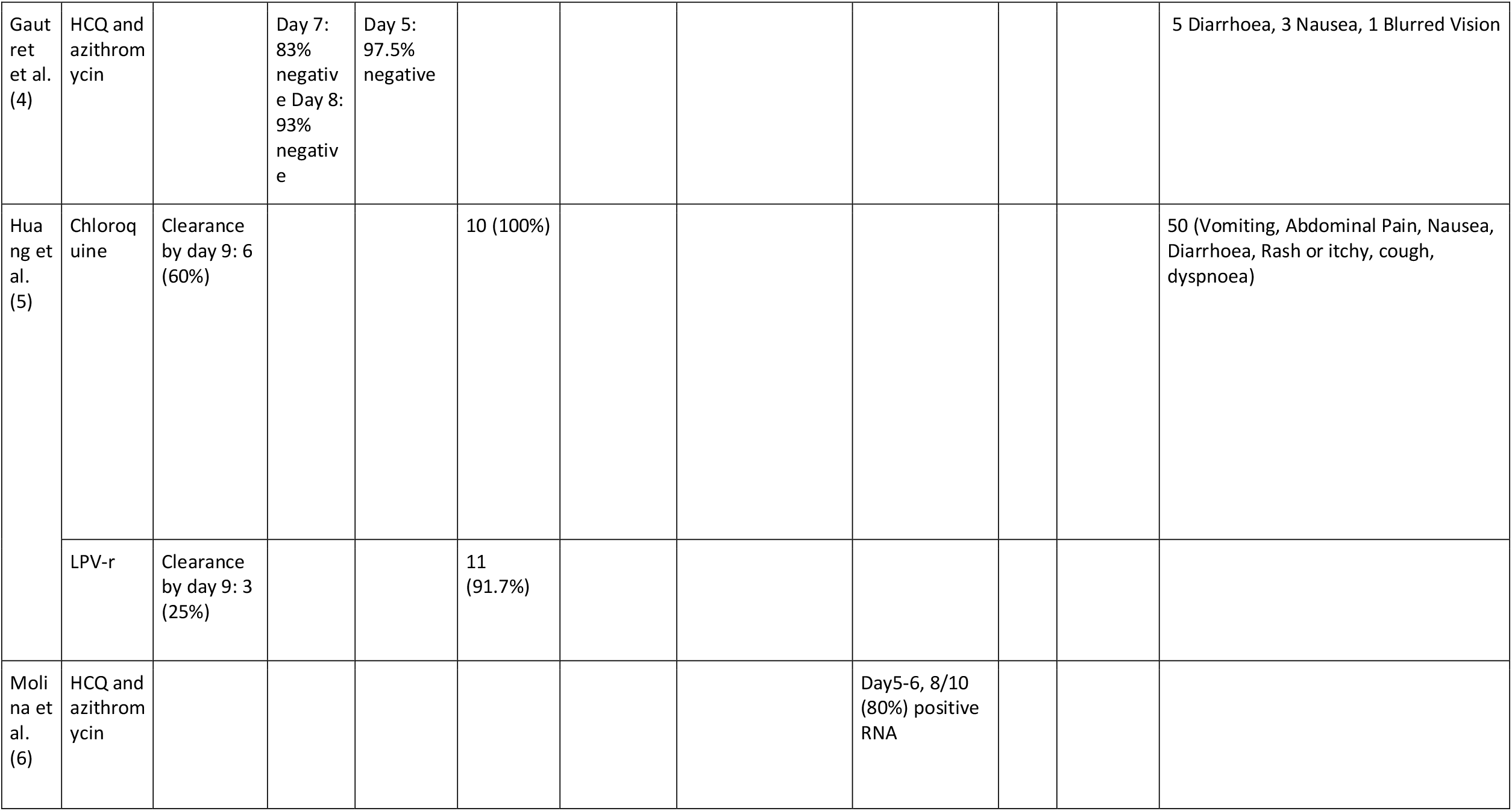
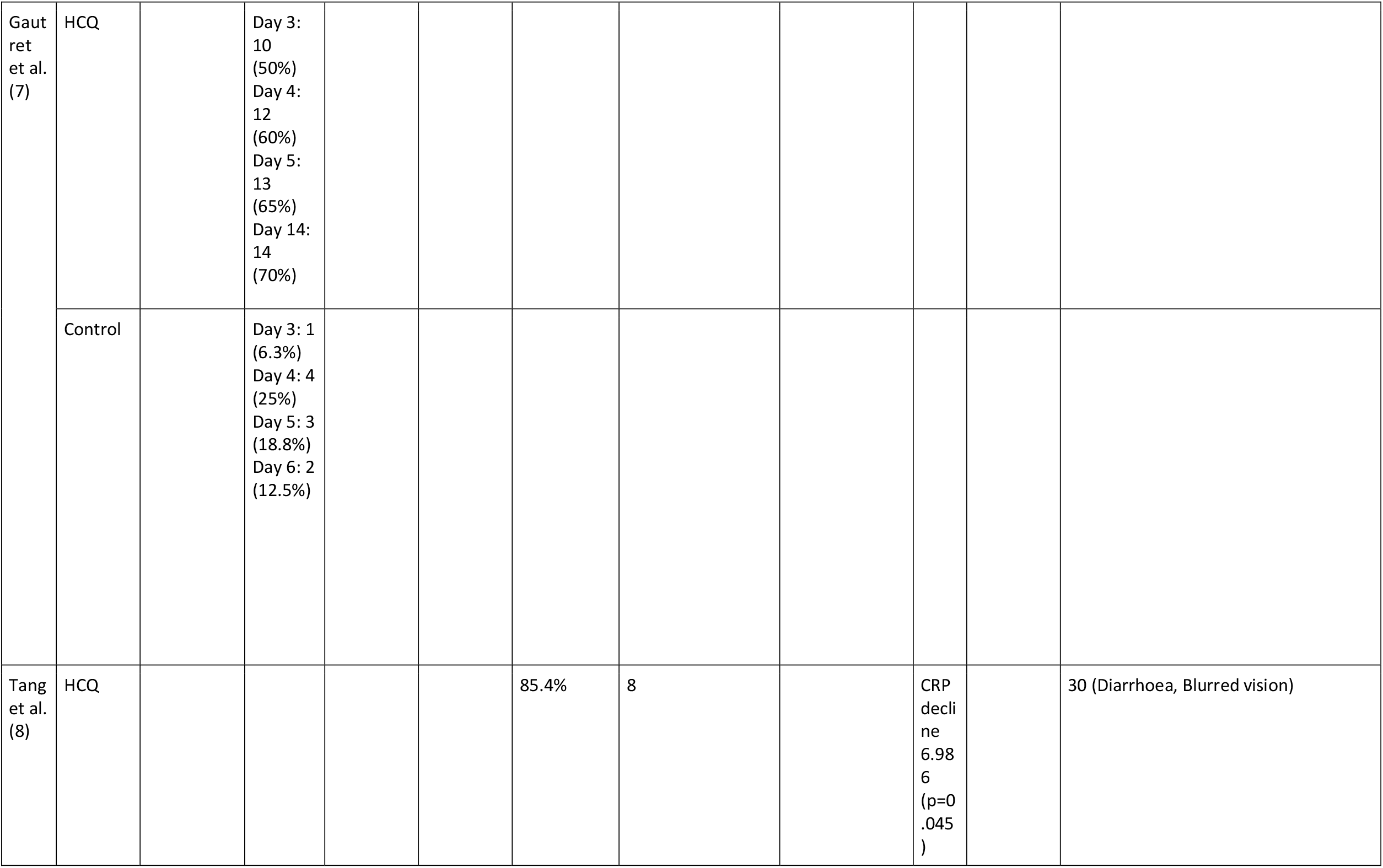
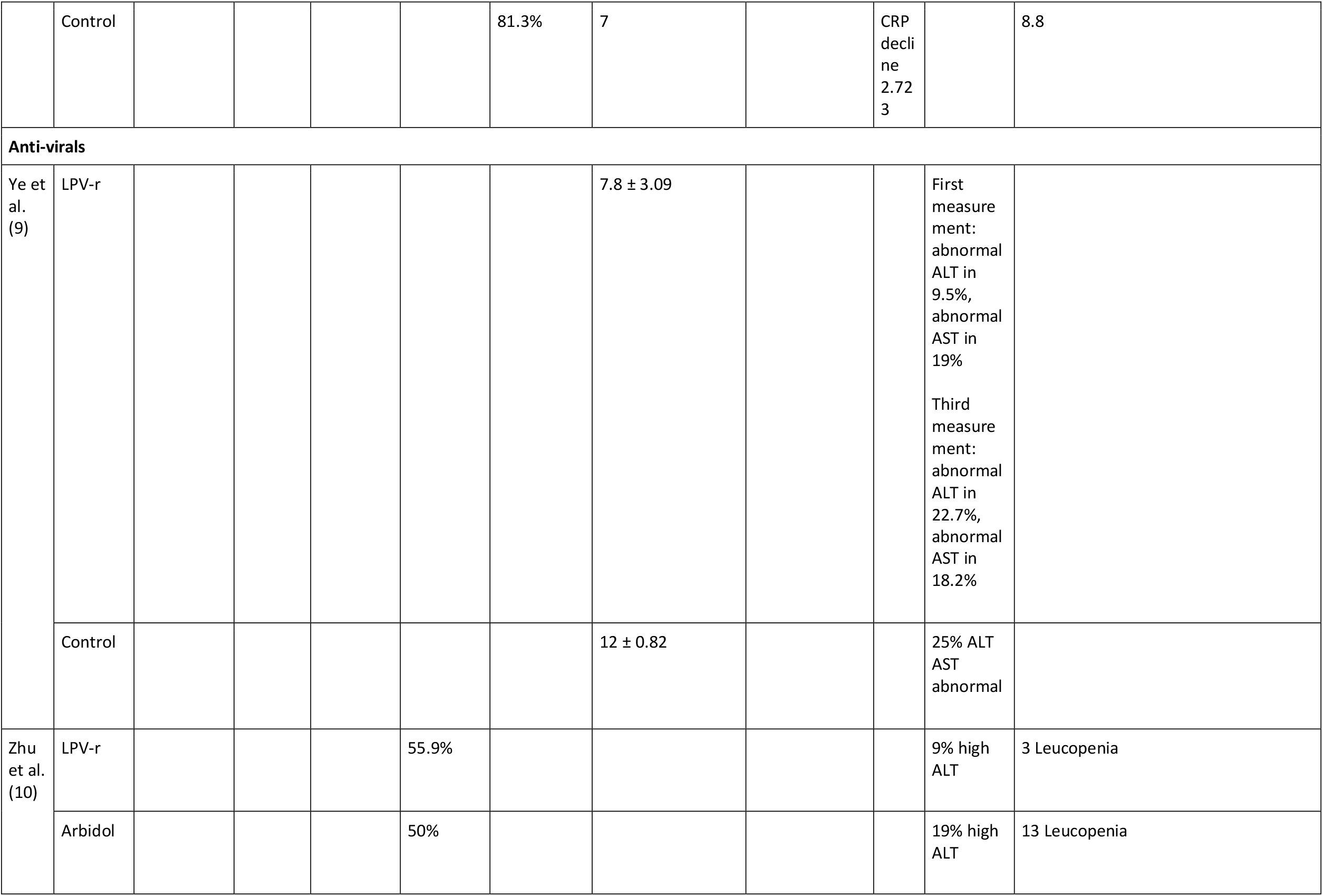
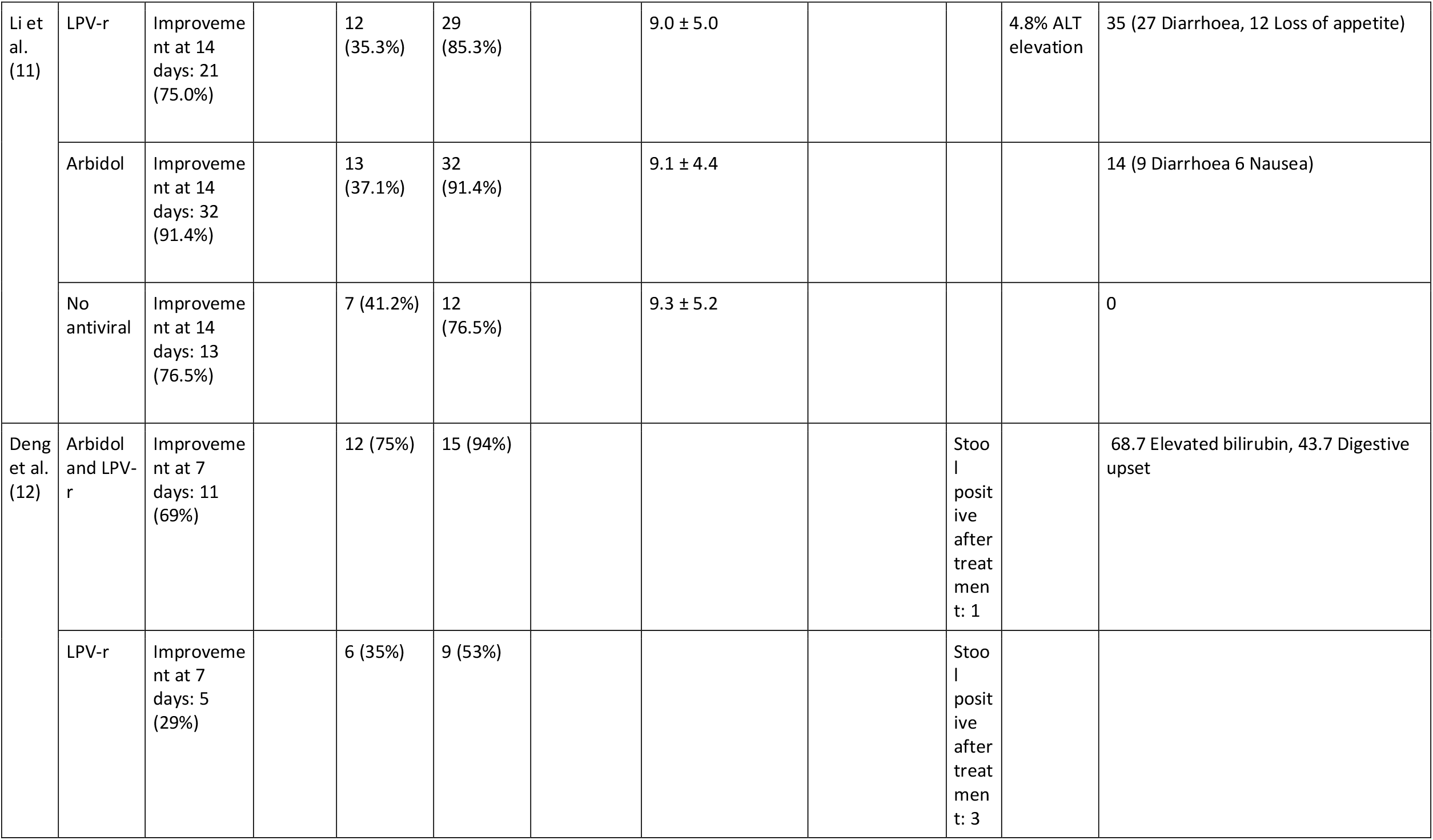
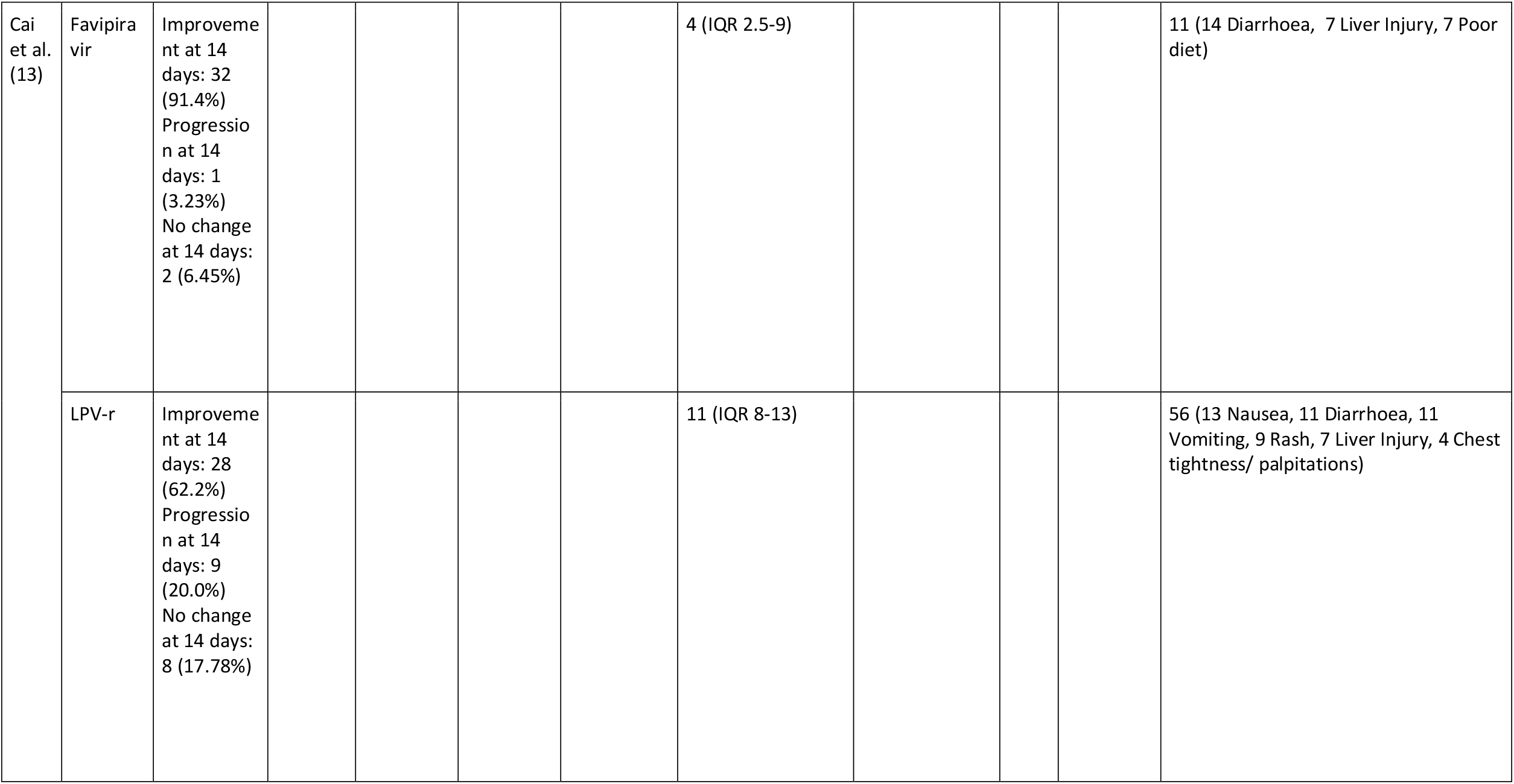
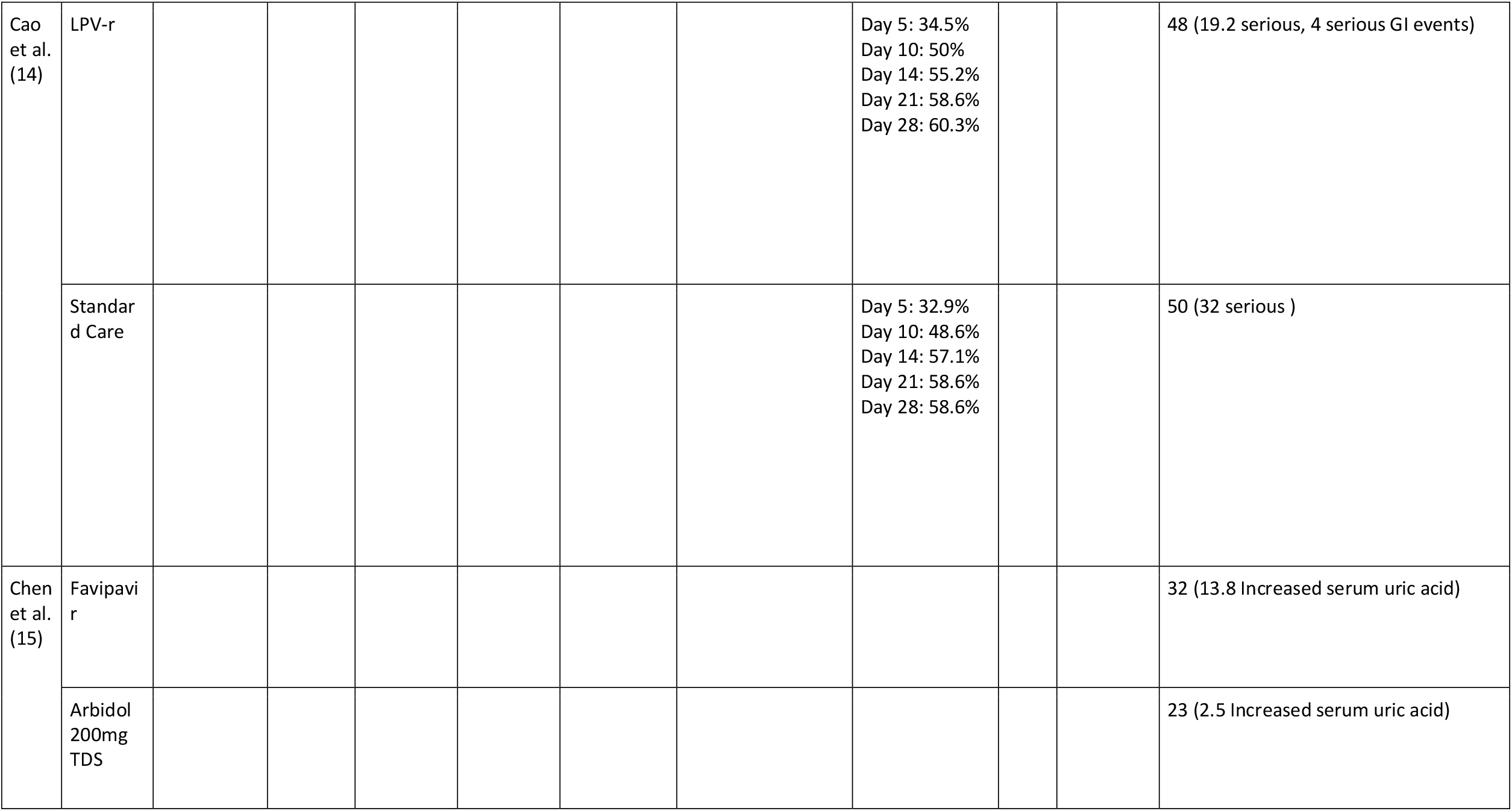
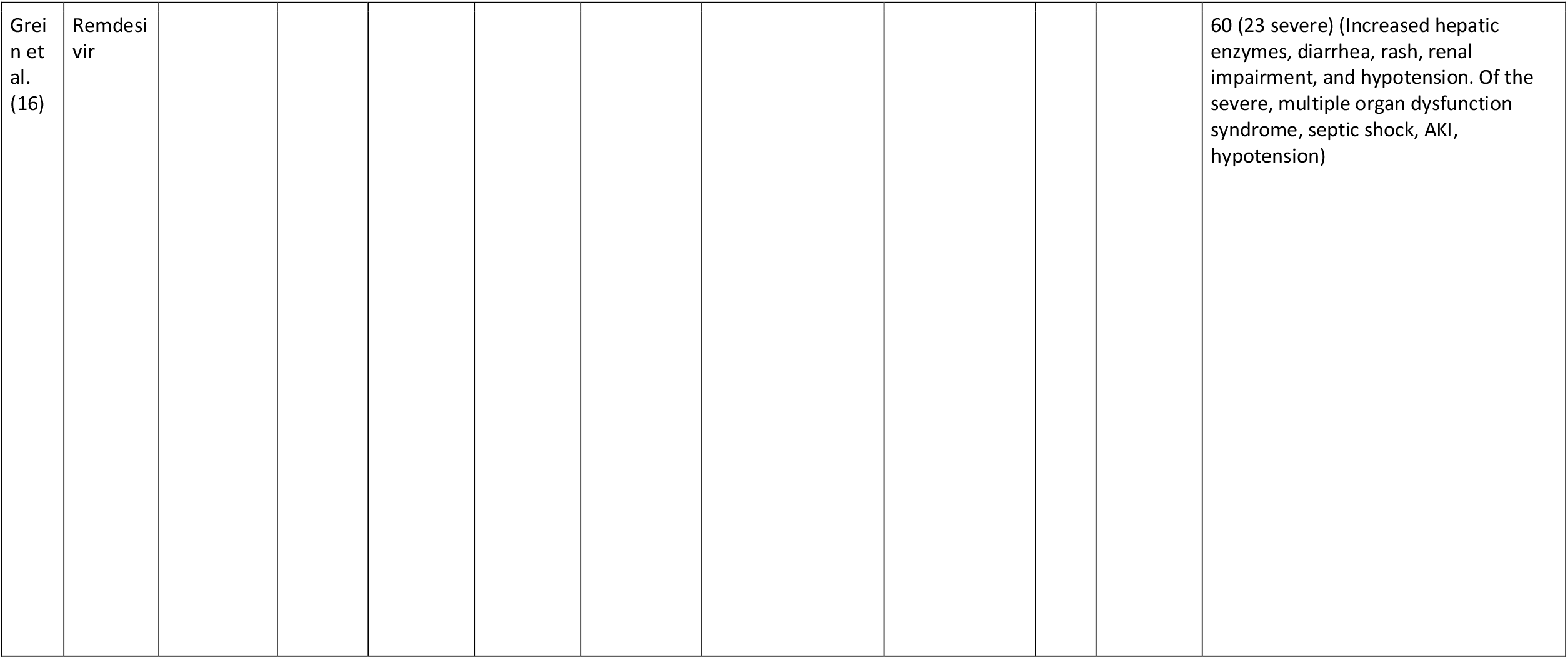
Radiology, Virology, Adverse Events

### Chloroquine (CQ)

2 studies examined the effects of CQ, 1 compared it to anti-viral treatment and 1 was a case series used in a news briefing. 1 study analysed both clinical recovery and viral clearance. CQ was initially mentioned in a news briefing (3), with results from more than 100 patients which had showed it was superior to the control treatment in shortening the disease course as well as inhibiting the exacerbation of pneumonia, improving lung imaging findings and promoting a virus negative conversion.

Subsequently, Huang et al. (5) compared it to another proposed therapy (LPV-r) in a cohort study with 10 patients in the CQ arm and 12 patients in the lopinavir/ritonavir arm. With CQ, 70% of the patients had a negative reverse transcriptase-polymerase chain reaction (RT-PCR) at day 7, 90% at day 10 and 100% at day 14. Compared to the LPV-r group where 58% of the patients had a negative RT-PCR at day 7, 75% at day 10 and 92% at day 14. There was a delay in lung clearance based on CT imaging, suggesting viral clearance does not translate immediately into pathological improvement in lungs. With CQ, 20% had a CT scan improvement at day 10 and 100% at day 14 compared to anti-virals with 8% showing an improvement at day 10 and 75% at day 14. With CQ, discharge rates from hospital were 100% at day 14 but with anti-virals, only 50% of the patients were discharged at day 14.

### Hydroxychloroquine (HCQ)

6 studies examined the effects of HCQ, of which 2 also added azithromycin. 4 compared it to a standardized care and 2 were case series with HCQ and azithromycin. 4 studies analysed clinical recovery, whilst 6 studied viral clearance

There have been 3 RCTs in China. In the study by Chen et al. (1), where the eligibility criteria are not known, 30 patients were recruited, with 15 patients in each arm. It showed no significant difference (p>0.05) in negative seroconversion rate at day 7 with 87% negative in the HCQ group and 93% in the control group and in time to negative seroconversion (p>0.05). Time to normalization of body temperature was also comparable between the 2 arms and radiological progression using CT imaging showed 33% of the cases progressed of the HCQ group compared to 47% in the control group.

Chen et al. (2) has included only mild cases with oxygen saturation (SaO2) >93% and confirmed chest CT with pneumonia, comparing HCQ (n=31) to standard treatment (n=31) which was undefined. The primary outcome was time to clinical recovery, defined as cough relief and return of body temperature (<36.6°C on surface, <37.2°C under armpit and mouth or <37.8°C in rectum and tympanic membrane) maintained for more than 72 hours. It was shown that normalization of body temperature was significantly (p=0.0008) shorter by 1 day in the HCQ group (2.2 days) compared to the control group (3.2 days). There was also significantly (p=0.0016) reduced cough remission time in the HCQ group (2 days) compared to the control group (3.1 days). It was also noted that overall 6% of the patients progressed to a severe illness and these occurred in the control group not receiving HCQ.

The largest randomized controlled trial (8) recruited a total of 150 patients enrolled from 16 centres, 75 in the HCQ arm and 75 in the standard of care (SOC) arm which included provision of intravenous fluids, supplemental oxygen, regulatory laboratory tests, SARS-CoV-2 testing, haemodynamic monitoring and intensive care. This study included anyone over the age of 18 with a confirmed SARS-CoV-2 infection. The primary outcome was the negative conversion of SARS-CoV-2 within 28 days. Secondary outcomes included alleviation of clinical symptoms defined as resolving fever to axillary temperature of <36.6°C, normalization of SaO2>94% on room air, disappearance of respiratory symptoms; laboratory parameters and chest radiology within 28 days. Viral clearance was similar in both arms with a negative seroconversion rate at 28 days of 85.4% and 81.3% in the HCQ arm and SOC arm respectively. The time to negative seroconversion was also similar with 8 days and 7 days in HCQ and SOC arms respectively. The overall rate of symptom alleviation within 28 days was also similar with HCQ (59.9%) and with SOC alone (66.6%). The median time to alleviation of clinical symptoms was also similar with 19 days in the HCQ arm and 21 days in the SOC arm. However, in a subgroup analysis when confounding effects of other anti-viral agents was removed, the efficacy of HCQ on the alleviation of symptoms is more evident (Hazard ratio, 8.83, 95%CI, 1.09-71.3). Changes in c-reactive protein (CRP) and lymphocyte count were also analysed with the HCQ arm showing a significantly greater decline (p=0.045) in CRP from baseline (6.986) compared to the SOC arm (2.723) and a greater elevation in lymphocyte count from baseline in the HCQ (0.062) compared to SOC (0.008) which was not significant (p=0.547)

A French cohort study (7) enrolled 20 patients with HCQ and 16 patients was a control, regardless of their clinical status. It has shown that HCQ can provide clearing of nasopharyngeal carriage in 50% of the patients by day 3 compared to 6.3% without it. The same research group have reported a case series (4) with 80 patients on a combination of azithromycin and HCQ. This showed negative virus cultures in 97.5% of those included at day 5 with only 15% requiring oxygen and 4% requiring ICU. However, these results were rapidly questioned by another study (6) who followed the same regime of azithromycin and HCQ but showed that at day 5, 8 out of 10 (80%) of the patients were still positive for Covid-19 RNA.

### Antivirals

A total of 8 studies regarding anti-virals was included, 6 studies examined the effects of LPV-r. Of these, 2 compared it to a standardized care, 1 compared it to favipavir, 1 compared it to the combination of LPV-r and arbidol, 1 compared it to arbidol and 1 compared it to both arbidol and a control group. 1 other study compared favipavir to arbidol and a case series regarding remdesivir is also included. 4 studies analysed clinical recovery, whilst 5 studied viral clearance.

There are 2 RCTs comparing LPV-r to control and one further compares it to arbidol. Cao et al. (14) is the largest RCT in this systematic review enrolling 199 patients with pneumonia confirmed by chest imaging, SaO2<94% while on breathing ambient air, 99 in the LPV-r group and 100 to the standard-care group. The primary outcome was the time to clinical improvement, defined as time from randomization to improvement of two points on a seven category ordinal scale or live discharge which was 16 days in both LPV-r and standard-care alone. In subgroups of treatment within 12 days or later treatment, there was no association with a shorter time to clinical improvement. Secondary outcomes measured included a 28 day mortality which was lower in the LPV-r (19.2%) than in the standard-care (25% group), stay in intensive care which was shorter in the LPV-r (6 days) group than in the standard-care (11 days) group, percentage of patients with clinical improvement at day 14 which was higher in the LPV-r group (45.5%) than in the standard-care group (30%) and time to discharge which was shorter in the LPV-r group (12 days) than in the standard-care group (14 days). Other secondary outcomes included duration of oxygen therapy, duration of hospitalization and time from randomization to death which had no significant difference between the two groups. In terms of virology, the percentage of patients with detectable viral RNA was similar in both groups on any sampling day (5, 10, 14, 21 and 28), for example at day 5, 34% in LPVr group compared to 32.9% in standard-care group. However, the mortality in this trial was 22.1% which is significantly higher than the mortality reported in descriptive studies, potentially indicating that a severely ill population was recruited. Therefore, the question of whether this anti-viral treatment may be effective in early treatment remains unaddressed.

Another RCT (11) recruiting patients with mild (mild symptoms but no signs of pneumonia on imaging) or moderate (fever, respiratory symptoms and pneumonia on imaging) Covid-19 compared LPV-r, arbidol and a control group with no anti-viral medication. The primary outcome was time of positive-to-negative conversion of SARS-CoV-2 from initiation of treatment to day 21 which was not significantly different (p=0.981). Time to viral clearance was 9 days, 9.1 days and 9.3 days in LPV-r, arbidol and control group respectively. There was also no significant difference of negative conversion rates at day 7 (p=0.966) and day 14 (p=0.352) of treatment. There was also no significant difference in secondary outcomes which included rate of antipyresis, rate of cough resolution and rate of improvement of chest CT imaging at day 7 and 14 (p>0.05). To take into account the influence of time from onset to treatment, this was evaluated in those who deteriorate to a severe clinical status (5 days) compared to those who did not deteriorate (4 days) and showed no significant difference (p=0.619).

However, one cohort study (9) comparing 42 patients treated with LPV-r to 5 patients in the control group has shown a significant reduction in time for body temperature normalization, 4.8 days in the LPV-r group compared to 7.3 days in the control group (p=0.0364).

In one study (12), LPV-r has also been used with arbidol which has higher negative seroconversion rate both at day 7 with 75% negative in the combination group compared to 35% in the LPV-r group (p<0.05) and at day 14, 94% negative in the combination group compared to 53% negative in the LPV-r group (p<0.05). There was also a significant difference (p<0.05) in the chest CT scans showing improvement in the combination group (69%) when compared to the LPV-r group (29%).

Another cohort study (15) has suggested favipavir is superior with a higher clinical recovery rate at day 7, defined as >72h recovery of body (axillary) temperature <36.6°C, respiratory rate, SaO2 >98% and cough relief (mild or none). This was significantly different (p<0.0001) with 61% of the patients clinically recovered in the favipavir group, compared to the 52% in the LPV-r group.

A case series (16) involving Remdesivir for compassionate use in 53 patients showed a 68% of the patients improved in the category of oxygen support by day 18 with 15% of the patients worsening. The cumulative incidence of clinical improvement, defined by either a decrease of 2 or more points on the six point ordinal scale or live discharge, was 84% improvement. In this case series, 7 (13%) of the patients died, 6 of which were receiving non-invasive ventilation. Overall mortality from date of admission was 0.56 per 100 hospitalisation days and when comparing patients receiving invasive ventilation (0.57) to those receiving non invasive ventilation (0.51), there was no substantial difference. However, the risk of death was greater among patients over the age of 70 (hazard ratio 11.34) and those with a higher serum creatinine (hazard ration 1.91).

Post-searching, a RCT done at 10 hospitals in Wuhan was published (31). This enrolled 237 patients who were Covid-19 positive, had pneumonia confirmed by chest imaging, had SaO2<94% on room air and were within 12 days of symptoms onset, 158 to the remdesivir arm and 79 to the placebo arm. However, only 155 and 78 in the remdesivir and placebo arm respectively were included in the per-protocol population due to withdrawal of consent, receiving the medication for less days than the protocol and not starting the study. The primary clinical endpoint was time to clinical improvement defined as a two-point reduction in patients’ admission status on a six-point scale. This was not significantly different in the remdesivir group (21 days) compared to the placebo group (23 days). In those receiving treatment within 10 days, there was a numerically faster time to clinical improvement in those in the remdesivir arm (18 days) compared to the placebo arm (23 days). Clinical improvement rates at day 7, 14 and day 28 were not significantly different between the remdesivir group and placebo group. However, numerically, at day 14 there was a higher clinical improvement rate at day 14 (27% with remdesivir compared to 23% in placebo) and at day 28 (65% with remdesivir compared to 58% in placebo). The 28 day mortality was similar between the remdesivir group (14%) and the placebo group (13%).

Other clinical outcomes such as duration of oxygen support, duration of invasive mechanical ventilation duration of hospital stay, time to discharge and time to death were not significantly different. However, numerically, the days of invasive mechanical ventilation were lower in the remdesivir group (7 days) compared to the placebo group (15.5 days). In terms of viral load, no differences were observed between both groups with a similar decrease in viral load. In the same study, adverse events were reported in 66% of the patients (18% serious) in the remdesivir group, the most common ones being constipation, hypoalbuminaemia, hypokalaemia, anaemia, thrombocytopaemia and increased total bilirubin. Adverse events were also reported in 64% (26% serious) of the patients in the control group, including hypoalbuminaemia, constipation, anaemia, hypokalaemia, increased aspartate aminotransferase, increased bloods lipids and increased total bilirubin. Overall, more patients discontinued the drug due to adverse events in the remdesivir group (12%) than in the placebo group (5%).

### Adverse events

Adverse events are summarized in Table 3. Adverse events relating to HCQ were reported in 3 studies with an average incidence of 21% (range 6-30), including symptoms of diarrhea, blurred vision, nausea, rash, headache and abnormal liver function tests. The adverse events of CQ were only reported in one study as 50% including vomiting, abdominal Pain, nausea, diarrhoea, rash, cough and dyspnea.

Antiviral side effects ranged from 0-60% across the studies with reported side effects such as liver injury (high transaminases, high bilirubin), leucopenia, gastro-intestinal and cutaneous side effects (diarrhea, vomiting, nausea, rash). One randomized controlled trial reported LPV-r adverse effects as 48% (19.2% serious ones) compared to 49% (32% serious ones) in the control group. The same study reported that 14% of the patients could not complete the 14 day course of LPV-r due to adverse events. There was only one case series regarding remdesivir which reported a 60% adverse event rate, including increased hepatic enzymes, renal impairment, diarrhea, rash and hypotension. This included 23% of severe adverse events including multiple organ dysfunction syndrome, septic shock, acute kidney injury and hypotension.

### Case Reports

16 case reports (32-46) are summarised in Table 4. In each of the cases, a number of medications have been trialed, antibiotics in 10 of the cases, HCQ in 8, LPV-r in 7 and remdesivir in 2 of the cases. Interestingly, only 2 of them reported adverse effects, both with the use of HCQ and azithromycin: high transaminases, Atrial Fibrillation and long QT syndrome.

**Table 4:** Case Reports

| Author | Title | Age | Gender | Comorbidities | Medication given | Clinical improvement from admission | Adverse events |
| --- | --- | --- | --- | --- | --- | --- | --- |
| Cheng et al. (32) | First case of Coronavirus Disease 2019 (Covid-19) pneumonia in Taiwan | 55 | F | Hypothyroidism | Antibiotics - ceftriacone, amoxicillin/clavulanate | Day 27: oxygen therapy ceased<br>Day 28: discharged |  |
| Falcão et al. (33) | Case Report: Hepatotoxicity Associated with the Use of Hydroxychloroquine in a Patient with Novel Coronavirus Disease (Covid-19) | 29 | F | Recent childbirth post-partum haemorrhage | Azithromycin, Piperacillin tazobactam, Hydroxychloroquine | Day 16: remained in ICU but clinically improving | High transaminases |
| Gabriels et al. (34) | Inpatient use of mobile continuous telemetry for Covid-19 patients treated with hydroxychloroquine and azithromycin | 72 | F | Paroxysmal atrial fibrillation | Hydroxychloroquine and azithromycin |  | AF, long QTc |
| Guilpain et al. (35) | Rituximab for granulomatosis with polyangiitis in the pandemic of Covid-19: lessons from a case with severe pneumonia | 52 | F | Granulomatosis with polyangiitis, overweight (BMI 27), hypertension | Lopinavir/ ritonavir for 3 days from Day12.<br>Hydroxychloroquine for 10 days from | Day 20: extubated<br>Day 25: oxygen therapy ceased<br>Day 29: seronegative and discharged | None reported |
| Han et al. (36) | The course of clinical diagnosis and treatment of a case infected with coronavirus disease 2019 | M | F | Hypertension, type 2 diabetes, 20-year smoker | Lopinavir and ritonavir, methylprednisolone, recombinant human interferon alfa-2b, ambroxol hydrochloride, moxifloxacin hydrochloride | Day 10: discharged |  |
| Hillaker et al. (37) | Delayed Initiation of Remdesivir in a COVID-19 Positive Patient | 40 | M | Anxiety, depression, obese (BMI 31), hypercholesterolaemia, 5-year smoker | Hydroxychloroquine 5 days, Remdesivir at day9 813 days after onset) | Day 12: extubated<br>Day 13: oxygen therapy ceased |  |
| Holshue et al. (38) | First Case of 2019 Novel Coronavirus in the United States | 35 | M | Hypertriglycercaemia | Vancomycin and cefepime. Day6 (day 10 illness), IV remdesivir | Day 8: oxygen therapy ceased | None reported |
| Kim et al. (39) | The First Case of 2019 Novel Coronavirus Pneumonia Imported into Korea from Wuhan, China: Implication for Infection Prevention and Control Measures | 35 | F | Obese (BMI 33) | Lopinavir/ ritonavir | Day 11: resolution of fever | None reported |
| Lim et al. (40) | Case of the Index Patient Who Caused Tertiary Transmission of Coronavirus Disease 2019 in Korea: the Application of Lopinavir/Ritonavir for the Treatment of Covid-19 Pneumonia Monitored by Quantitative RT-PCR | 54 | M | None | Lopinavir/ ritonavir from day8 | Day 9: resolution of fever |  |
| Liu et al. (41) | A Locally Transmitted Case of SARS-CoV-2 Infection in Taiwan | 52 | F | Type 2 diabetes | Oseltamivir and levofloxacin on day3 | Not reported |  |
| Mathies et al. (42) | A Case of SARS-CoV-2-pneumonia with successful antiviral therapy in a 77-year-old male with heart transplant | 77 | M | Heart transplant in 2003 (ischaemic cardiomyopathy), previous CMV-colitis and pneumonia, chronic kidney disease, hypertension, type 2 diabetes | Hydroxychloroquine and piperacillin/tazobactam and ganciclovir (Hx of CMV infections) | Day 7: oxygen therapy ceased<br>Day 12: discharged |  |
| Ning et al. (43) | Novel Coronavirus (SARS-CoV-2) Infection in A Renal Transplant Recipient: Case Report | 29 | M | Renal transplant in 2018 (chronic kidney disease), hypertension | Moxifloxacin, Trimethoprim, sulfamethoxazole. On day2, lopinavir/ritonavir. Antibacterial stopped. | Day 13: discharged |  |
| Song et al. (44) | Coronavirus Disease 19 (Covid-19) complicated with pneumonia in a patient with rheumatoid arthritis receiving conventional disease-modifying antirheumatic drugs | 61 | F | Rheumatoid arthritis | Lopinavir/ritonavir, regular medication - hydroxychloroquine, meloxicam and famotidine | Day24: discharged |  |
| Spezzani et al. (45) | Benign Covid-19 in an immunocompromised cancer patient – the case of a married couple | 60 | F | Metastatic breast cancer (chemotherapy-treated), colonic angiodysplasia, bicuspid aortic valve, lower limb varicosities | Levofloxacin, piperacillin/tazobactam, darunavir/ cobicistat, hydroxychloroquine | Day 6: discharged |  |
|  |  | 60 | M (husband) | Hypertension | Darunavir/cobicistat and hydroxychloroquine and ceftriaxone, azithromycin | Day 11: extubated<br>Day 23: discharged |  |
| Tang et al. (46) | Coronavirus Disease 2019 (COVID-19) Pneumonia in a Hemodialysis Patient | 50s | M | Renal failure (receiving haemodialysis), diabetes, hypertension, chronic hepatitis B | Moxifloxacin and lopinavir/ ritonavir | Day 8: discharged |  |

### Risk of Bias

Figure 2 and 3 show the risk of bias assessment of each study, using RoB-2 for RCTs and ROBINS-I for non-RCTs. Of the 6 RCTs, 1 (1) was of high risk whilst 5 (11, 12, 14, 16) were of moderate risk. Of the non RCTs, 1 was not appropriate for analysis (15), 1 (13) was of moderate risk and 8 (4-7, 9, 10, 12, 16) were of serious risk.

### Meta analysis

3 studies were included in the meta-analysis of adverse events using HCQ (Figure 4) and 2 studies were included in the meta-analysis of negative seroconversion rate using HCQ (Figure 5). In the meta-analysis of seroconversion rate, Cheng et al. measured this rate at day 7 while Tang et al. measured it at day 28. These show that there is a significant difference (p=0.001) regarding adverse events in HCQ compared to control group (Odd ratio 3.61, 95% CI, 1.66-7.84). However, no difference was found (p=0.68) in negative seroconversion rate between HCQ and the control group (Odds ratio 1.18, 95% CI, 0.53-2.66)

**Figure 4:**
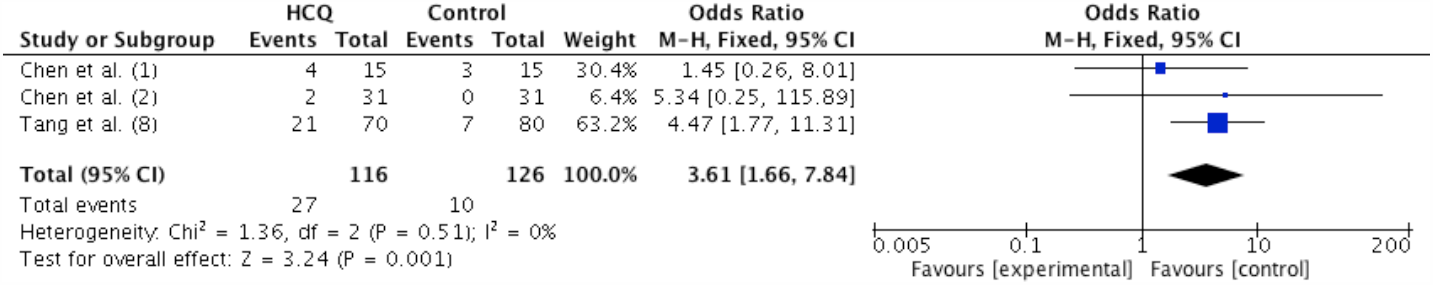
Effect of HCQ compared to control group on adverse events.

**Figure 5:**
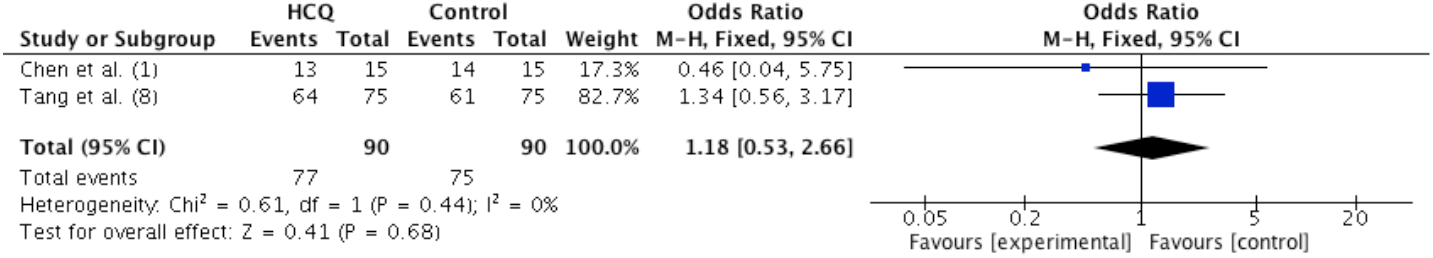
Effect of HCQ compared to control group on negative seroconversion rate.

3 studies were included in the meta-analyses of adverse events with LPV-r (Figure 6). This shows that there is no significant difference (p=0.1) between LPV-r and control group regarding adverse events (Odds ratio 1.54, 95% CI, 0.92-2.55).

**Figure 6:**
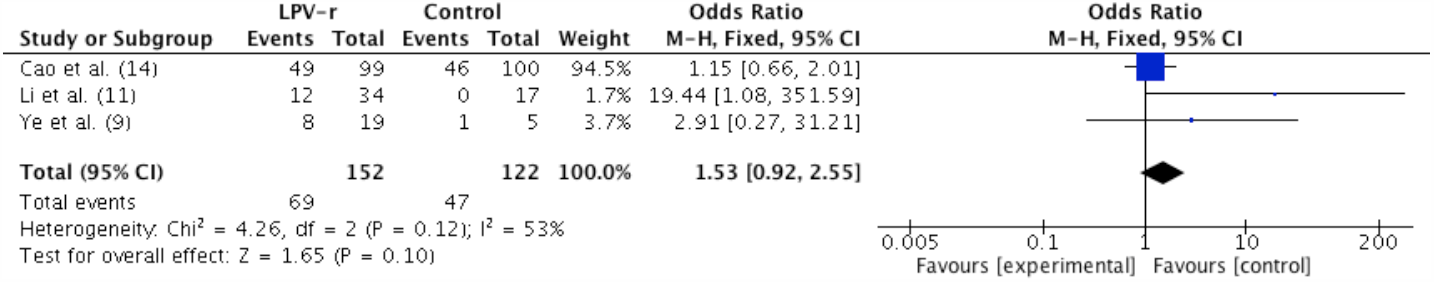
Effect of Lopinavir-ritonavir compared to control group on adverse events.

## Discussion

Having been declared a global pandemic, with the numbers of cases rising everyday, Covid-19 has been an unprecedented challenge in a number of areas with a subsequent reorganization of the clinical activities (47). Clinicians across the world have trialed repurposing a number of medications. Regarding CQ, it has been suggested as a more effective and inexpensive option when compared to anti-virals (5). A recent trial (27) has compared the use of high-dosage (12g) and low-dosage CQ and has suggested that a high-dosage regimen with azithromycin and oseltamivir was not safe to continue due to concerns regarding cardiotoxic events, myocarditis and QTc interval prolongation which can be associated with an increase in fatal arrhythmias such as ventricular tachycardia. However, age can be a confounder as it can be associated with unfavourable outcomes. Therefore, it was advised that high dosage CQ should not be recommended in the treatment of severe Covid-19 but these findings cannot be extrapolated to patients with non-severe Covid-19.

The results from randomized controlled trials using HCQ have shown a similar negative seroconversion rate and time to clinical recovery, when compared to standard care. However, it has been recorded that HCQ may fasten normalization of body temperature and cough remission. With some smaller cohorts showing benefit of HCQ, it is important to be cautious with results. Therefore, overall, the role of of HCQ in the management for Covid-19 may still remain promising but larger scale studies are required.

Anti-virals are another group of medications which have been investigated with both RCTs showing no difference when compared to standard-care alone. However, one RCT has shown that LPV-r may have a lower 28-day mortality, a shorter stay in intensive care, a shorter time to discharge and an effect on clinical improvement. The only study (16) regarding remdesivir identified in our search was an observational study about a compassionate use in patients with severe Covid-19, which showed an 84% clinical improvement and that improved in the category of oxygen support in 68% of the cases. After our search was carried out, a RCT (31) was published comparing remdesivir and placebo, showing no significant advantage in time to clinical improvement, mortality or time to viral clearance with remdesivir, even if well tolerated. However, the power of this study was insufficient as it did not reach its target enrolment, due to the marked reductions in new patient presentations in mid-March in Wuhan. The initiation of treatment might have been quite late in the disease course as there were restrictions on hospital bed availability to have a significant improvement in outcomes.

When summarizing the case reports, it is challenging to draw conclusions; however, the clinical improvement of the patients reported with a use of a cocktail of medications is an important aspect to consider.

The present systematic review has several limitations. The paucity of RCTs, the ‘gold standard’ for comparing interventions, is probably due to difficulties with randomization and blinding in the unprecedented and stressful environment faced by healthcare services. Therefore, a large number of studies are observational or cohort studies which are quicker to organize and implement, therefore obtaining results rapidly which is essential as the number of worldwide cases is rising at an alarming rate. Another limitation is that studies vary in the outcomes measured, therefore side-by-side comparisons become more difficult. Due to this heterogeneity of the studies, we were only able to perform a meta-analysis on a maximum of 3 studies. In one of the analyses regarding negative seroconversion rate, they were measured at different time points, therefore not giving an accurate view. A large proportion of the studies included also do not measure survival and morbidity outcomes which are important. Many of the studies included have also not been formally peer reviewed yet but, due to the urgency of the pandemic, draft manuscripts have been uploaded.

We included every study found in our systematic review which may have introduced bias, therefore it is important to analyse all studies cautiously as the selection of patients is important as different studies have recruited patients with a different severity of Covid-19. It is also important to consider the timeline of the administration of the medication as early administration of medication could be more beneficial than later in the course of the disease. When using risk of bias assessment tools, most of the non-RCTs were at serious risk of bias due to confounders including baseline confounding factors such as the presence of comorbidities which could affect the outcome of the patient. No blinding of patients or assessors can also pose a risk of bias due to the potential unreliable measurement of outcomes.

To our knowledge, this is the first systematic review regarding the repurposing of drugs in Covid-19, summarizing 16 studies as well as 16 case reports. In conclusion, it is difficult to establish the efficacy of these drugs in the management of Covid-19 due to the lack of evidence. However, likewise, there is also no sufficient evidence to show these drugs are not successful in improving clinical outcomes and reducing viral load. Therefore, it is important to balance the benefits of trialing this medication with the adverse events described where the spectrum of these has not been clearly understood.

Accordingly, there is a need for further high quality data, especially from RCTs, to evaluate the benefits of these repurposed drugs in the treatment of patients with Covid-19. We recognise there is a large global effort for this, with the biggest Covid-19 trial, the RECOVERY trial (48), having recruited over 8500 participants across 173 sites to date (1 May 2020). This randomized trial is inviting clinically suspected or laboratory confirmed Covid-19 adult (>18 years) patients hospitalised in the United Kingdom to participate with randomisation to one of 4 arms: usual care, usual care plus LPV-r, usual care plus low-dose dexamethasone, usual care plus HCQ or usual care plus azithromycin. Those who deteriorate are further randomized between tocilizumab and a control group.

## Conclusion

This study indicates no clinical effectiveness regarding the role of chloroquine, hydroxychloroquine and anti-viral for the treatment of Covid-19 patients. However, there is potential for these medications but further large clinical trials are required to achieve more reliable findings. Therefore, a risk-benefit analysis is required on an individual basis to weigh out the potential improvement in clinical outcome and viral load reduction compared to the risks of the adverse events.

## Data Availability

All the data included are available

## Author contributions

All authors contributed to the planning, conduct and reporting of the work. All authors approved the submitted version

